# Multi-Analyte Liquid Biopsies for Molecular Pathway Guided Personalized Treatment Selection in Advanced Refractory Cancers: A Clinical Utility Pilot Study

**DOI:** 10.1101/2021.12.23.21268152

**Authors:** Darshana Patil, Dadasaheb Akolkar, Rajnish Nagarkar, Navin Srivastava, Vineet Datta, Sanket Patil, Sachin Apurwa, Ajay Srinivasan, Rajan Datar

**Author notes:** **Corresponding Author** Ajay Srinivasan, Department of Research and Innovations, Datar Cancer Genetics (Pvt.) Ltd., F-8 D Road, MIDC Ambad, Nasik, Maharashtra – 422010. India., 0091-253-6690803.

## Abstract

**Purpose:** The selection of safe and efficacious anticancer regimens for treatment of broadly refractory patients with metastatic cancers remains a clinical challenge. Such patients are often fatigued by toxicities of prior failed treatments and may have no further viable standard of care treatment options. Liquid Biopsy-based multi-analyte profiling in peripheral blood can identify a majority of drug targets that can guide the selection of efficacious combination regimens.

**Patients and Methods:** LIQUID IMPACT was a pilot clinical study where patients with advanced refractory cancers received combination anticancer treatment regimens based on multi-analyte liquid biopsy (MLB) profiling of circulating tumor biomarkers; this study design was based on the findings of prior feasibility analysis to determine the abundance of targetable variants in blood specimens from 1299 real-world cases of advanced refractory cancers.

**Results:** Among the 29 patients in the intent to treat (ITT) cohort of the trial, 26 were finally evaluable as per study criteria out of whom 12 patients showed Partial Response (PR) indicating an Objective Response Rate (ORR) of 46.2% and 11 patients showed Stable Disease (SD) indicating the Disease Control Rate (DCR) to be 88.5%. The median Progression-Free Survival (mPFS) and median Overall Survival (mOS) were 4.3 months (95% CI: 3.0 – 5.6 months) and 8.8 months (95% CI: 7.0 – 10.7 months), respectively. Toxicities were manageable and there were no treatment-related deaths.

**Conclusion:** The study findings suggest that MLB could be used to assist treatment selection in heavily pretreated patients with advanced refractory cancers.

## Introduction

Clinical management of patients with advanced broadly refractory cancers faces challenges due to unavailability of standard of care systemic anticancer regimens as well as adverse impact on health due to accumulated toxicities of prior failed treatments. The National Comprehensive Cancer Network (NCCN) guidelines for several cancers, hence, often recommend palliative care or clinical trial in such cases. We have previously reported findings from the RESILIENT trial which showed that de novo multi-analyte profiling of tumor tissue and tumor derived component in blood can inform selection of safe and efficacious treatments even in heavily pre-treated advanced refractory cancer cases ^1^. The benefit of such treatments could not be extended to several patients who were screened for inclusion in the RESILIENT trial, but excluded since they were unable to undergo an invasive biopsy due to anatomical constraints (proximity of the lesion to vital organs) or co-morbidities ^2, 3^. In order to address this challenge, we designed a Multi-analyte Liquid Biopsy (MLB) which evaluated circulating tumor analytes in peripheral blood to determine targetable (molecular and functional) features and guide selection of patient-specific label-agnostic combination anticancer regimens. The rationale for combination regimens rather than monotherapies was based on the hypothesis that combination regimens can be potentially beneficial in targeting multiple vulnerabilities and / or blocking escape or resistance mechanisms. The design of MLB combined the strengths of a multi-analyte tumor profiling with the convenience and safety of a liquid biopsy. The MLB evaluated cell free DNA (cfDNA) and circulating tumor associated cells (CTACs). ctDNA was profiled for gene variants including gain of copy, point mutations and indels.C-TACs were profiled by immunocytochemistry (ICC) to determine overexpression of therapeutic target proteins as well as by in vitro chemo resistance / chemo response profiling against a panel of FDA approved anticancer agents. We first ascertained feasibility of MLB in an analysis of 1299 patient specimens, where circulating tumor analytes were profiled for the molecular and functional features detailed above. This feasibility analysis helped estimate the proportion of patients where an actionable indication may be observed. Subsequently, we conducted LIQUID IMPACT, a pilot basket trial which evaluated and established the clinical utility of MLB to inform selection of safe and efficacious patient-specific combination regimens in a cohort of heavily pre-treated advanced refractory cancers. Study participants provided blood specimens which were evaluated to select personalized combination regimens with targeted and cytotoxic anticancer agents. Patients were assigned to treatment baskets based on targetable molecular indication(s) and the performance of each basket was evaluated in addition to the overall performance to determine suitability for a future study with an expanded cohort.

## Methods

### Study Design – Feasibility Study

For the feasibility study cohort of 1299 cancer patients, peripheral blood specimens from the study sponsor’s (Datar Cancer Genetics, DCG) test services and prior research studies were utilized. Patients who had availed of the sponsor’s test services, as well as all research study participants, had previously provided written informed consent approved by Ethics Committees of the study sponsor, for research use of specimens and de-identified data. The feasibility study was based on analysis of existing (leftover) specimens and specimen data and did not involve the collection of new specimens.

### Study Design –Prospective Study

LIQUID IMPACT (Trial Registration number CTRI/2019/02/017548, http://ctri.nic.in/Clinicaltrials/pmaindet2.php?trialid=31265) was a pilot prospective, single arm, non-randomized interventional study for evaluation of MLB-guided personalized combination treatment regimens in patients with advanced refractory solid organ cancers. The trial was approved by the Institutional Review Boards (IRB) and Ethics Committees of the Study Sponsor as well as the trial site (HCG-Manavata Cancer Centre, HCG-MCC). The trial was conducted in accordance with all applicable ethical guidelines and the Declaration of Helsinki. The primary outcome measure of the trial was objective response rate (ORR) defined as the proportion of participants where radiological complete response (CR) or partial response (PR) was observed as per RECIST 1.1 criteria. Secondary outcome measures included clinical benefit rate (CBR), i.e. the proportion of participants with CR, PR or stable disease (SD) for ≥ 60 days, Progression Free Survival (PFS), Overall Survival (OS) and Quality of Life (QoL). In addition to treatment response, safety was also an important consideration since participants were being administered combination regimens of anticancer agents in a label-agnostic setting. The single-arm design of the trial was based on earlier RESILIENT [1], NCI-MATCH and ASCO TAPUR basket trials where the pilot phase did not include a control arm ^4–6^. Considering the diversity in cancer types and unique treatment history of each patient, the trial acknowledged that there can be no accurate external control for each patient.

### Statistical Methods and Analysis

The sample size of the prospective clinical study was determined based on the ORR, assuming that the ORR in such refractory advanced stage cancer patients is <10%. Simon’s 2-stage design was used to validate the adequacy of cohort size for assessment of MLB based therapy. The null hypothesis that the true response rate is 10% was tested against a one-sided alternative. Initially, at least 10 patients were required to accrue; if there was 1 or no response, the study was to be terminated. Otherwise, at least 15 additional patients were required to accrue. The null hypothesis would be rejected if 5 or more responses were observed in 25 patients. With 25 evaluable patients, this design yields a type I error rate of 5% and power of 86% when the true response rate is 20%. The 95% CI of ORR was constructed using a binomial distribution (Clopper-Pearson estimation method). Patient demographics were analysed with descriptive statistics. Contingency tables described the categorical data with counts and percentages. Continuous data was summarized using median and range. CONSORT diagram, Waterfall Plot and Bar Graphs were used to summarize the data. Kaplan-Meier estimator was used to estimate survival function.

### Feasibility Study Cohort

The feasibility cohort consisted of 1299 cancer patients across different cancer types, such as breast (n = 304), colorectal (n = 167), non-small cell lung cancer (n = 130), pancreas (n = 94), ovary (n = 88), sarcoma (n = 61), prostate (n = 60), head and neck (n = 49), esophagus (n = 43), melanoma (n = 33) and central nervous system (n = 32) (Refer Supplementary Table S1 for details). cfDNA analysis by Next-Generation Sequencing (NGS), gene expression profiling by immunocytochemistry (ICC) and in vitro chemo response profiling on C-TACs was performed on 1299 cancer specimens.

### Prospective Study Cohort

The LIQUID IMPACT prospective trial recruited patients with refractory solid organ cancers who had either failed at least two prior lines of systemic Standard of Care (SoC) anticancer treatments or where (further) systemic SoC treatment options were unavailable / unviable, and where an invasive biopsy to obtain tumor tissue (for *de novo* tumor profiling) was not possible. Eligible patients had radiologically measurable lesions with an Eastern Co-operative Oncology Group (ECOG) performance status of ≤ 2 and fitness as ascertained by the treating clinician. Patients who fulfilled the above criteria were counselled regarding the potential benefits and risks of the trial. Thereafter, patients who provided signed informed consents were enrolled.

### Multi-Analyte Liquid Biopsy

All study participants provided only peripheral blood specimens. The process of multi-analyte liquid biopsy and formulation of patient-specific therapy recommendations (TR) has been described previously ^7, 8^. The complete details of investigations under multi-analyte liquid biopsy are provided in Supplementary Methods. The study did not evaluate PD-L1, Microsatellite Instability (MSI) or Mismatch Repair (MMR) status in patients since these evaluations required tumor tissue (which was neither available nor feasible to obtain in this cohort) and at the time of study recruitment there were no approved non-invasive tests to determine the status of these biomarkers in blood samples. Consequently, immune checkpoint inhibitors (ICIs) were not considered for inclusion in the treatment regimens.

### Treatments

In all patients in the prospective (LIQUID IMPACT) cohort, the treatment regimens were initially administered at lower (≤50%) doses, and gradually escalated based on close monitoring of toxicity. Patient-specific regimens were administered until either disease progression, death or dose limiting toxicity was encountered. Patients with disease progression were excluded from the trial and shifted to other SoC treatment (if available) or physician’s choice treatment or considered for best supportive care alone.

### Evaluations

Patients in the prospective study underwent an ^18^F-Fluorodeoxyglucose Positron Emission Tomography – Computed Tomography (FDG PET-CT) scan before initiation of treatment to determine baseline status of the disease. The response was evaluated as per RECIST 1.1 criteria ^9^ from follow-up scans following at least two treatment cycles or 60 days of treatment, except in cases where the treating clinician advised evaluation in the interim.

### Endpoints

The primary endpoint of the prospective pilot study was Objective Response Rate (ORR) defined as the percentage of patients who achieved Complete Response (CR) or Partial Response (PR) during the active study phase. Progression Free Survival (PFS) was also evaluated in patients and was defined as time from commencement of treatment under ETA (Encyclopedic Tumor Analysis) to disease progression or death during the active study phase. Quality of Life (QoL) was evaluated based on patient’s feedback on symptomatic and functional status at baseline and at study termination or most recently available follow-up.

### Patient Monitoring

All patients in the prospective study underwent periodic clinical evaluations to assess fitness for treatment as per study protocol. Adverse events (AEs) were recorded every week either during patient admissions or by telephonic follow-up. All AEs were reported as per NCI-CTCAE v5 criteria ^10^. Grade 3 and above AEs, if any, were followed up till resolution. Patients were followed up until study termination or patient exclusion (death / loss to follow-up / withdrawal of consent), whichever was earlier to determine Progression Free Survival (PFS).

## Results

### Feasibility Study

Specimens from different cancer patients availing the study sponsor’s test services along with specimens from prior research studies conducted by the study sponsor (n = 1299) were evaluated for the feasibility of multi-analyte liquid biopsy, patient stratification and selection of tailored anticancer treatment regimens. The dendrogram in Supplementary Figure S1 provides details of specimens in the feasibility study cohort, sourced from the test service and research study arms, respectively.

### Molecular Landscape of the Feasibility Study Cohort

Figure 1 illustrates the snapshot of molecular features in the feasibility study cohort. In the analysis of 1299 cancer patients, we identified molecular/functional features in 807 (62.12%) patients. TP53 mutations were seen in 43% of specimens, KRAS mutations in 12% of specimens and NRAS, GNAS and MYC mutations were found in <3% of specimens each. Copy Number Variations (CNV) as gain of copy were observed frequently in CCND, CDK, MYC, MET and FGFR. The multigene panel does not provide information on loss of gene copy. Overexpression of EGFR, VEGF, VEGFR and mTOR proteins as determined by ICC were seen in 52-62% of specimens indicating the possibility of selection of respective targeted therapies against EGFR, angiogenesis and mTOR pathways. Among other angiogenesis pathway targets, PDGFR and FGFR based indications for anti-angiogenesis agents were observed in 1% and 4% of specimens respectively while further indications for targeting mTOR pathway based on variants in PIK3CA were detected in 11% of specimens. Targetable indications in HER2 (ERBB) and ER (ESR) were identified in 4% and 3% of specimens.

**Figure 1.**
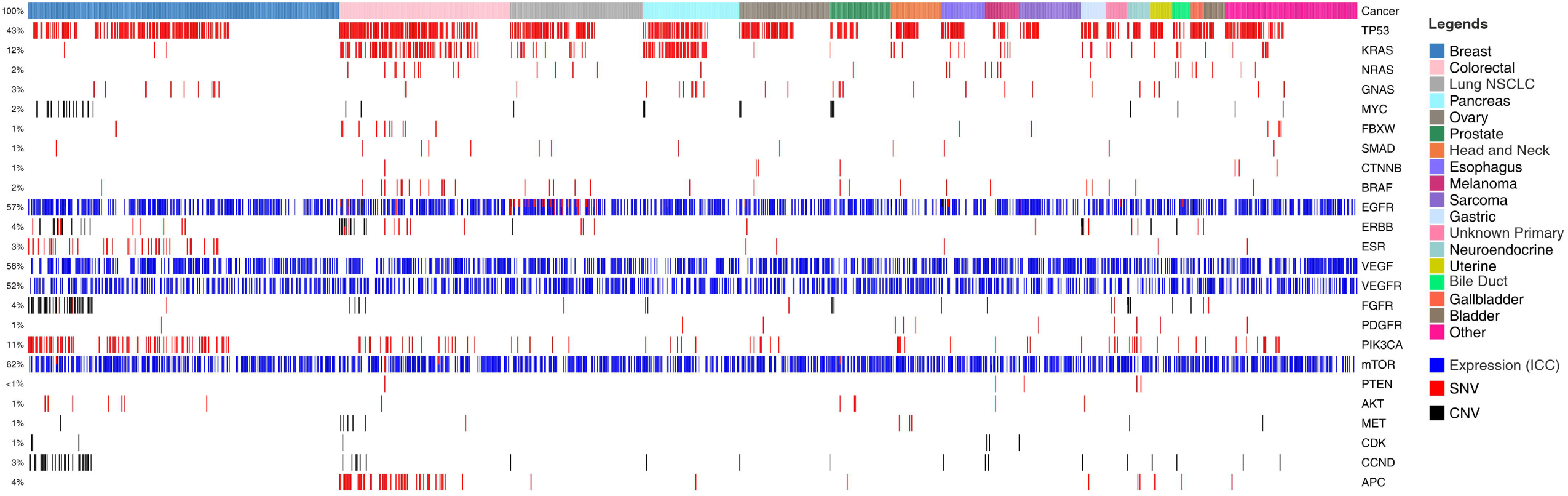
Landscape of Genomic Alterations in the Feasibility Study Cohort. Colour coded boxes in the topmost row indicate cancer types. Each vertical column indicates a single patient. Features such as Single Nucleotide Variations (SNVs) are represented by red colour; Copy Number Variations (CNVs; gain) by black; and ICC based gene expression analysis by blue. Gene names are enlisted on right Y-axis and the % of specimens harbouring the different features are enlisted on left Y-axis.

In vitro chemo response profiling data was also available for 1299 specimens where the C-TACs were treated with a panel of 30 anticancer drugs and the response determined. The panel included anticancer agents administered as SoC options (NCCN guidelines) as well as those which were ‘off-label’ drugs (non-SoC options). Among the 1299 specimens, C-TACs in 1087 (84%) showed response (cytotoxicity) to at least 1 SoC drug, C-TACs in 827 samples (64%) showed response to 2 or more drugs and C-TACs in 598 specimens (46%) showed response to 3 or more drugs. Interestingly, when the C-TACs were assessed for response to off-label (non-SoC) drugs, these numbers were 91% (≥ 1 drug), 80% (≥ 2 drugs) and 69% (≥ 3 drugs) respectively. The analysis thus makes available previously unexplored potential treatment options for heavily pretreated cancer patients where SoC options are either unavailable or unviable. The cancer-wise details of these sensitivities are presented in Supplementary Table S2, while Supplementary Figure S2 shows the % of specimens per cancer type which were responsive to each of the 30 anticancer agents tested. These findings suggested that treatment with either of these anticancer agents in a label agnostic manner may have potential clinical benefit.

### Prospective Study

For the prospective LIQUID IMPACT study cohort, 64 patients were screened between Feb 2019 and Oct 2019, of whom 56 were recruited, 43 were enrolled (received MLB guided therapy recommendations) and 29 patients eventually started treatment as per MLB. Thirty five patients were excluded between screening and start of treatment for various reasons including non-measurable lesions (n = 3), unfavourable or deteriorating Eastern Co-operative Oncology Group Performance Score (ECOG PS, n = 14), withdrawal of consent (n = 5) or absence of therapeutically targetable molecular indications (n = 13). Out of the 29 patients who started treatment, 3 were excluded within the first week (prior to any follow-up evaluation) for either ECOG PS deterioration (n = 1), withdrawal of consent (n = 1) and loss to follow up (n = 1), respectively. Twenty-six patients received MLB guided treatments and were evaluable as per study criteria. The CONSORT diagram (Figure 2) depicts the study structure and patient flow. Patient demographics, baseline status and prior treatments are provided in Supplementary Tables S3, S4, and S5 respectively.

**Figure 2.**
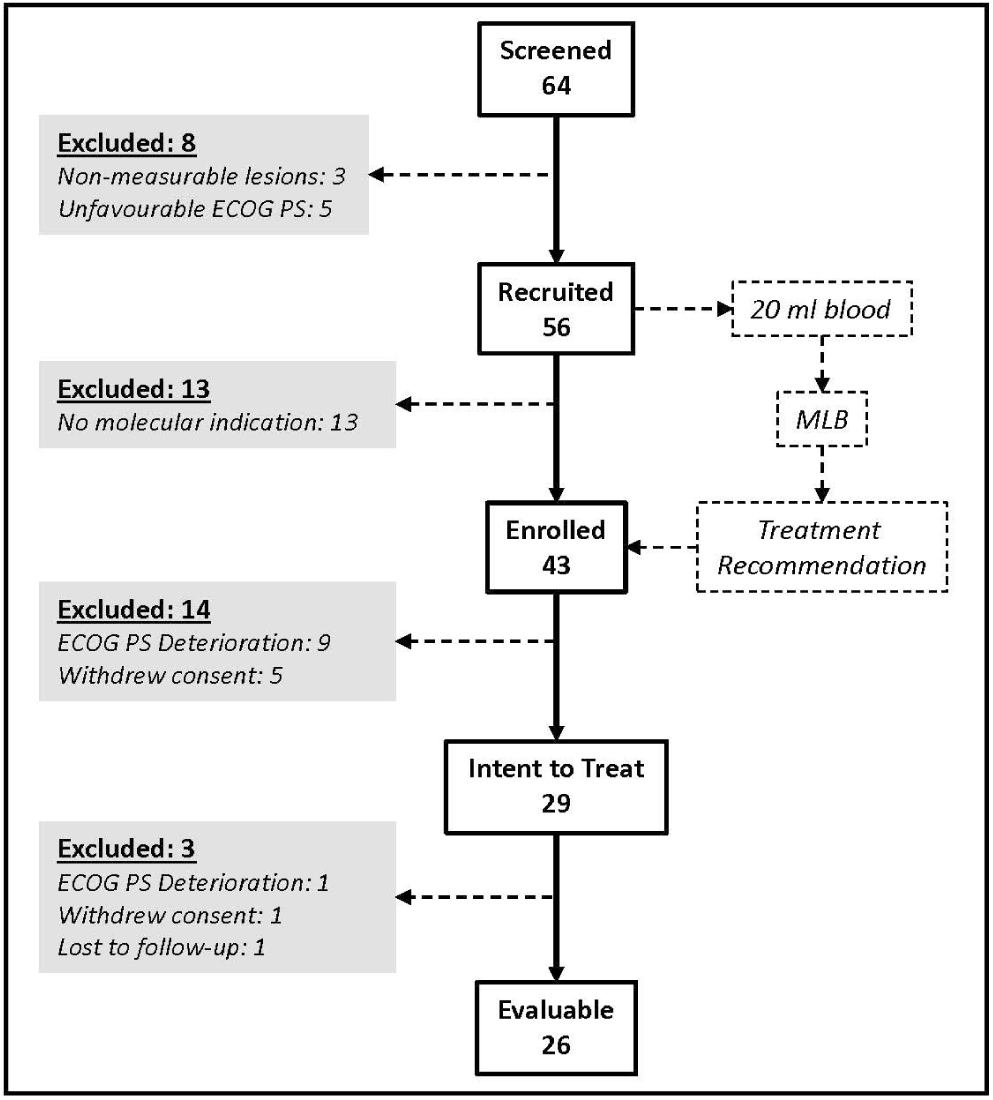
LIQUID IMPACT CONSORT Diagram. Among the 64 patients who were screened, 29 eventually started treatment (ITT population) and 26 were finally evaluable based on study criteria. Patients provided blood specimens at baseline which was used for multi-analyte liquid biopsy.

### Molecular Landscape of the Prospective ITT Cohort

The landscape of molecular features in the prospective Intent to Treat (ITT) population is depicted in Figure 3. Single nucleotide variations (SNV) in TP53 were a frequently encountered (11, 37.9%) feature followed by those in PIK3CA (4, 13.8%) and KRAS/NRAS (2, 6.9%). Among the ITT population, actionable SNVs were detected in 9 patients, of whom 8 were evaluable; actionable CNVs were detected in 2 patients, all of whom were evaluable, and gene overexpression was detected by ICC in 22 patients, of whom 19 were evaluable. Patient-wise actionable gene alterations that formed the basis for therapy selection are indicated in Supplementary Dataset.

**Figure 3.**
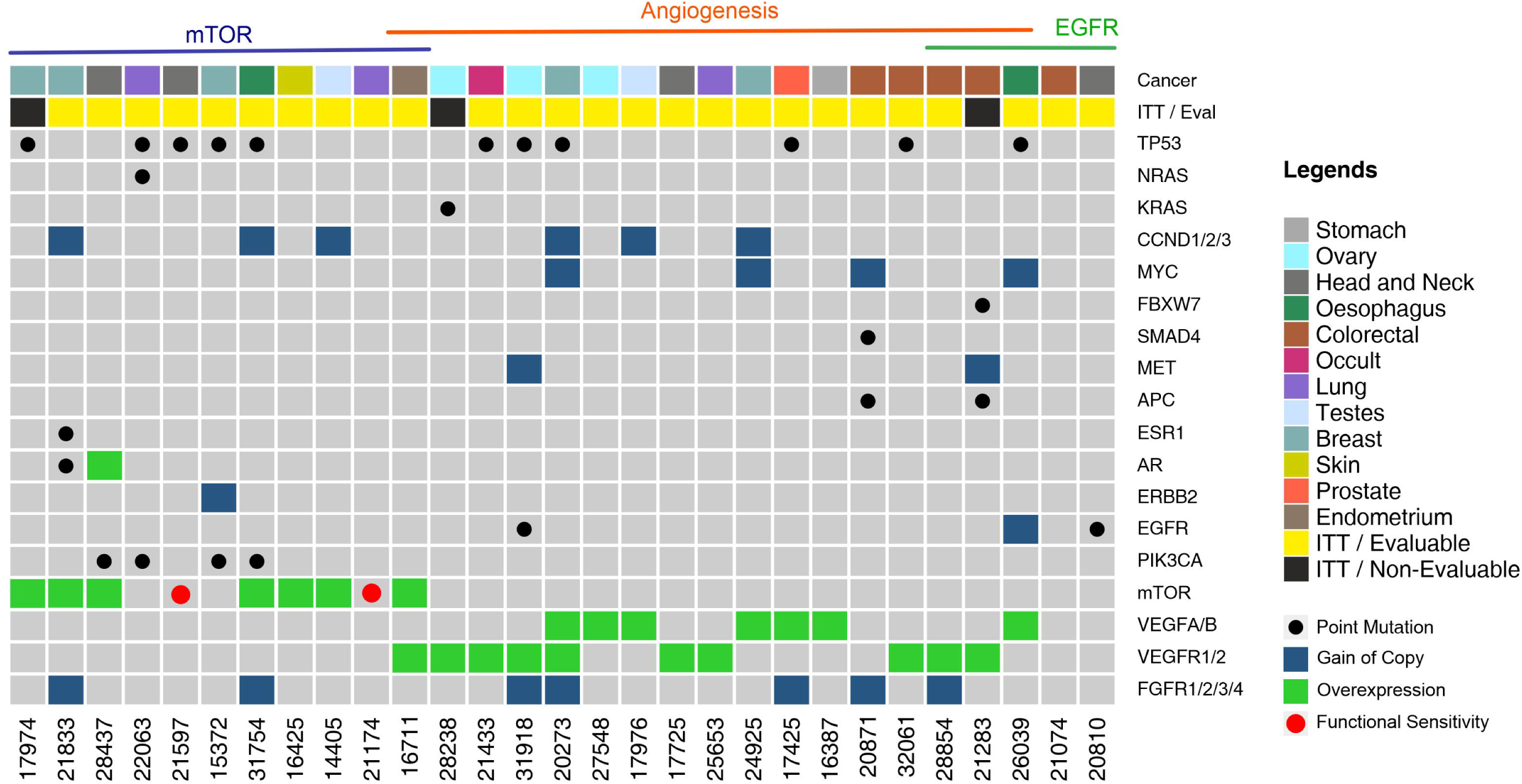
Landscape of Genomic Alterations in the Prospective Intent to Treat (ITT) Population. Each vertical column indicates a single patient (patient number in the bottom X-axis). Vertically stacked grey boxes in each column indicate individual genes (gene names on right Y-axis). Black dots within each box indicates a point mutation (single nucleotide variation), red dots indicate in vitro sensitivity (chemo response profiling, CRP), blue shaded boxes indicate gain of gene copy (CNV gain), green shaded boxes indicate gene overexpression. Patients are grouped by observed targetable molecular indications. Colour coded boxes in the topmost row indicate cancer types.

### Prospective Study Baskets and Treatments

Twenty-six of the initial 29 patients who received MLB guided treatments were evaluable for response as per study criteria. Among 11 patients (ITT) who were assigned an mTOR inhibitor, 10 were eventually evaluable. Among 17 patients (ITT) who were assigned an angiogenesis inhibitor, 15 were evaluable. Finally, among 5 patients (ITT) who received an EGFR inhibitor based on EGFR / ERBB2 activation or KRAS wild type, 4 were evaluable. One patient received an mTOR inhibitor as well as an angiogenesis inhibitor and was evaluated in both treatment baskets. Among 3 patients who received angiogenesis inhibitor as well as EGFR inhibitor, 2 were evaluable and were evaluated in both respective treatment baskets. Endocrine therapy was administered to 2 patients in addition to cytotoxic and targeted agents. Details of cancer types, treatment baskets and treatments are provided in Supplementary Table S6. Patient-wise details of prior treatments, MLB-indications and treatments are provided in Supplementary Dataset.

### Response to Treatment

Partial Response (PR) was observed in 12 of 26 evaluable patients yielding an ORR of 46.2% (41.4% in the ITT population of 29). Waterfall Charts depict the best response (Figure 4) of all 26 patients. Patients in the evaluable subset were followed up for a median duration of 4.6 months (range: 1.3 months – 13.2 months). PRs were observed in 5 (50%) of 10 evaluable patients who received an mTOR inhibitor, 8 (53.3%) of 15 evaluable patients who received an angiogenesis inhibitor and 2 (50%) of 4 evaluable patients who received an EGFR inhibitor. Though not an endpoint of the study, Stable Disease (SD) for more than 60 days was observed in 11 of the 26 overall evaluable patients, yielding a Disease Control Rate (DCR) of 88.5% in the evaluable population.

**Figure 4.**
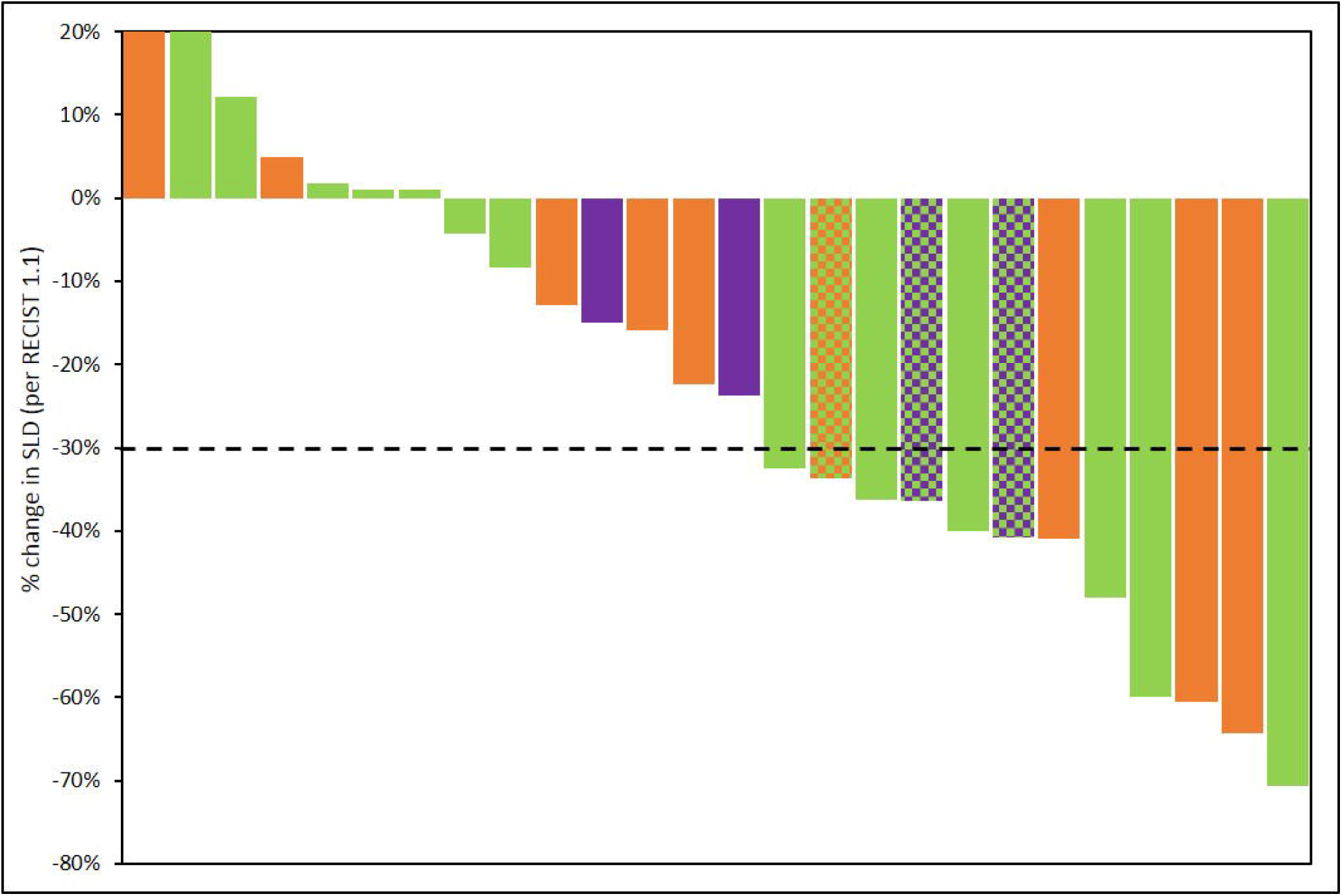
Summary of Outcomes. Treatment Response was evaluated as per RECIST 1.1. Percent change in dimensions of target lesions (Sum of Largest Diameters, SLD) between baseline and at best response are represented. Each bar represents the response in a unique patient. Patients are arranged in descending order of change (%) in SLD. Bars are colour coded as per the molecular indication: Orange: mTOR; Green: Angiogenesis; Purple: EGFR. In 3 patients, 2 signalling pathways were detected which are represented by a pattern of 2-colours, each corresponding to the respective pathway.

Median Progression Free Survival (mPFS) in the evaluable subset was 4.3 months (95% CI: 3.0 – 5.6 months). Similarly, mPFS was 3.8 months among 10 patients with mTOR activation, 5.2 months among 15 patients with angiogenesis activation and 7.0 months among 4 patients with EGFR activation. While true median Overall Survival (mOS) is indeterminate (several patients were lost to follow-up due to Covid pandemic), considering OS data censored at the most recent follow-up (Oct 2020), the mOS in the entire evaluable cohort was 8.8 months (95% CI: 7.0 – 10.7 months), 10.7 months among 10 patients with mTOR activation, 8.8 months among 15 patients with angiogenesis activation, 9.5 months among 4 patients with EGFR activation. The Kaplan Meier plots of PFS and OS are depicted in Figure 5. Details of overall response and per study basket are provided in Supplementary Table S7.

**Figure 5.**
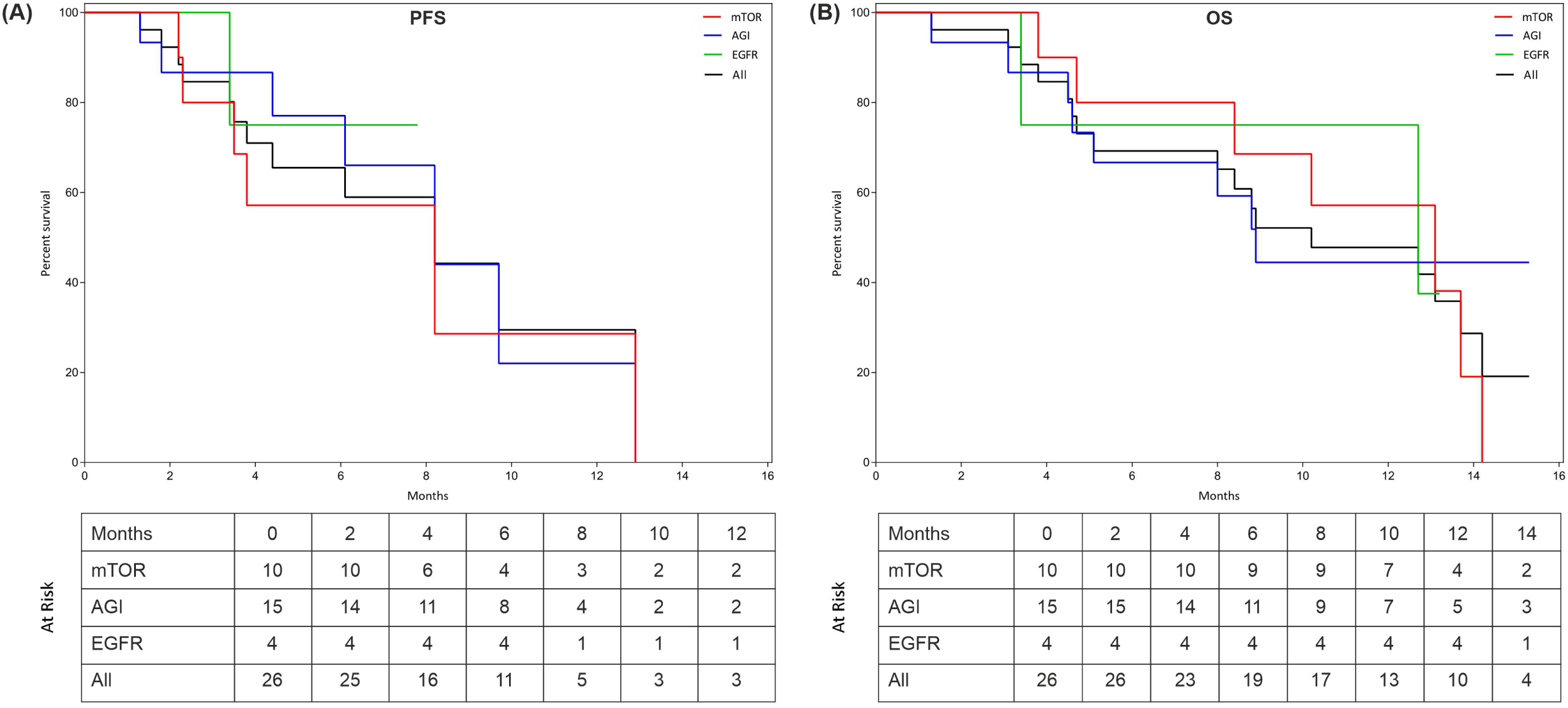
Progression Free Survival (PFS) and Overall Survival (OS). The Kaplan Meier plots show PFS (A) and OS (B) in all patients as well as in sub-cohorts where activation of EGFR, Angiogenesis (AGI) or mTOR signalling was observed. Patients at risk at each milestone are indicated in the inset table. Censoring events are indicated by a vertical cross bar.

### Adverse Events

All the 29 patients in the Intent to Treat (ITT) population were evaluated for Therapy Related Adverse Events (TR-AE). TR-AEs of any grade were reported in 25 (86.2%) of the overall population whereas Grade III and IV AEs were observed in 15 (51.7%) and 3 (10.3%) of the ITT cohort. Grade III TR-AEs which were considered as SAEs were observed in 34.5% of the overall population. All AEs were transient, acute (non-chronic), managed by standard procedures and followed up until resolution; Most Grade IV AEs and Grade III SAEs were resolved between 48-72 hours. There were no treatment related deaths. Overall and basket-wise profiles of AEs are provided in Supplementary Table S8.

### Quality of Life (QoL)

Quality of Life was measured based on the ECOG Performance Status as well as a brief questionnaire which evaluated the patients’ symptomatic status, ability to perform activities of daily living (ADL) and the ability to perform additional / more strenuous activities. Patients’ feedback was obtained at baseline and every month until study completion or exclusion. Within the evaluable cohort, 96% of patients indicated stable to decreased symptomatic/ECOG status while 46% of patients indicated stable to improved functional/ADL status.

## Discussion

We report the clinical potential of MLB-guided treatment selection in a cohort of heavily pre-treated cases of advanced refractory cancers. The significant ORR of 46.2% and manageable toxicities suggest that multi-analyte evaluations can identify multiple actionable vulnerabilities of a cancer, which can be treated with combination regimens of targeted and cytotoxic anticancer agents. The response rates reported in this pilot study justify the need for a larger cohort. The outcomes in our study are clearly superior to the outcomes reported in prior studies which attempted to personalize anticancer treatments based on univariate molecular profiling of the tumor for selection of label-agnostic monotherapies. ^4, 5, 11–17^ The modest clinical benefits, if at all, reported by these prior trials highlight the limitations of their design.

MLB detected potentially targetable molecular indications in several patients, the abundance of these targets being generally concordant with what has been reported in other studies such as NCI-MATCH, SHIVA, MOSCATO and I-PREDICT. ^11, 12, 15, 17^

We show that in addition to gene variants (point mutations / copy number variations), MLB can provide information on upregulation of targetable pathways via ICC profiling of CTCs, akin to immunohistochemistry (IHC) on tumor tissue or cytology specimens. In a recent large (>30,000 specimens) cohort study, we have described the potential of ICC profiling of CTCs to provide diagnostically relevant information without requirement of tumor tissue ^8^. In the present paper, we describe the potential application of ICC towards theranostic guidance. While molecular indications form the basis for selection of targeted and endocrine agents, there are fewer biomarkers for selection of cytotoxic anticancer agents which remain the mainstay of treatments in several cancers. We have previously evaluated and described the accuracy and utility of C-TACs in more than 5000 patients from peripheral blood for in vitro chemo response profiling ^18^. In vitro chemo response profiling of viable tumor cells from a tissue biopsy has previously demonstrated limited potential ^19, 20^, and has been unviable due to the inability to obtain viable tumor cells for de novo analysis. In these regards, the analysis of CTCs and C-TACs has shown some promise ^18, 21–25^. In the present study, in vitro chemo response profiling of C-TACs was used to guide selection of cytotoxic anticancer agents which were used in conjunction with targeted anticancer agents. MLB encompasses clonal variations arising from tumor heterogeneity and also captures the profile of the ‘leading edge’ tumour cells, which may well be the most aggressive elements for metastatic spread and invasiveness ^26^.

Studies have shown that combination regimens yield improved benefits over monotherapy ^27, 28^. Illustratively, the combination of Everolimus and Lenvatinib has higher efficacy over Everolimus monotherapy in metastatic RCC ^29^. The combination of Alpelisib and Fulvestrant has yielded higher response rates (∼26%) in ER+/HER2-metastatic breast cancers than Alpelisib monotherapy ^30, 31^. It has been shown that monotherapy for blockade of VEGF / VEGFR signalling often yields transient responses followed by eventual resistance ^32, 33^, owing to which combination strategies to achieve tandem blockade of multiple signalling pathways have been proposed as a viable strategy ^34–37^.

Prior meta-analyses have shown that it is possible to safely administer *de novo* drug combinations in patients without increasing the risk of adverse events (AEs) ^38–40^. Where the cancer has progressed following failure of multiple prior systemic lines of therapy (as in the current study cohort), patients tend to be at an inherently higher risk of AEs due to cumulative toxicities from prior treatments. Hence, all patients were administered initial doses at ≤50% and with controlled dose escalation. This strategy effectively controlled AEs; while Grade III and IV AEs were observed, they were limited, transient and manageable. Significantly, there were no treatment related mortalities. This pilot study provides evidence of the efficacy and reasonable safety of patient specific label-agnostic combination anticancer regimens when selected on the basis of MLB. The study findings are encouraging and justify a future larger multi-arm trial with various molecular subtype baskets for treatment of broadly refractory cancers.

## Conclusions

The LIQUID IMPACT trial was intended to be a pilot study to evaluate MLB for its potential and clinical utility to inform selection of safe and efficacious patient-specific combination regimens in advanced refractory cancers. Owing to the pilot nature of the trial, the size of each basket as well as the entire cohort was limited and the study design excluded a traditional control arm; this design was based on the aims and design of the NCI-MATCH and ASCO TAPUR basket trials where the pilot phase did not include a control arm ^4–6^. The outcome of the study shows promise for a larger study with statistically relevant cohort size.

## Data Availability

All data pertaining to the studies described in this manuscript are provided in the manuscript and the supplementary material. There are no additional datasets.

## Abbreviations

ADL: Activities of Daily Living
C-ETACs: Circulating Ensembles of Tumor Associated Cells
CNV: Copy Number Variations
CR: Complete Response
C-TACs: Circulating Tumor Associated Cells
CTCs: Circulating Tumor Cells
DCR: Disease Control Rate
ECOG: Eastern Co-operative Oncology Group
EGFR: Epidermal Growth Factor Receptor
ER: Estrogen Receptor
ERBB2: Erb-B2 Receptor Tyrosine Kinase 2
ETA: Encyclopedic Tumor Analysis
FDA: Food and Drug Administration
FDG PET-CT: 18F-Fluorodeoxyglucose Positron Emission Tomography– Computed Tomography
HER2: Human Epidermal Growth Factor Receptor 2
HPE: Histopathological Evaluation
ICC: Immunocytochemistry
IHC: Immunohistochemistry
IRB: Institutional Review Boards
ITT: Intent To Treat
MLB: Multi-analyte Liquid Biopsy
mTOR: mechanistic Target Of Rapamycin
MTB: Multidisciplinary Tumor Board
NCI-CTCAE: National Cancer Institute-Common Terminology Criteria for Adverse Events
NCI-MATCH: National Cancer Institute-Molecular Analysis for Therapy Choice
NGS: Next-Generation Sequencing
ORR: Objective Response Rate
OS: Overall Survival
PET-CT: Positron Emission Tomography– Computed Tomography
PFS: Progression Free Survival
PIK3CA: Phosphatidylinositol-4, 5-Bisphosphate 3-Kinase Catalytic Subunit Alpha
PR: Partial Response
QoL: Quality of Life
RCC: Renal Cell Carcinoma
RECIST: Response Evaluation Criteria in Solid Tumors
SD: Stable Disease
SNV: Single Nucleotide Variations
SoC: Standard of Care
TAPUR: Targeted Agent and Profiling Utilization Registry
TR-AE: Therapy Related Adverse Events
VEGF: Vascular Endothelial Growth Factor
VEGFR: Vascular Endothelial Growth Factor Receptor

## Acknowledgements

The authors are grateful towards all study participants and their caregivers. The contributions of HCG Manavata Cancer Centre as well as the staff of the Study Sponsor (DCG) towards managing various clinical, operational and laboratory aspects of the study are also acknowledged with gratitude.

## Ethical Considerations

The Liquid Impact Prospective Study was approved by institutional review boards and ethics committees of the study sponsor (Datar Cancer Genetics, DCG) as well as clinical trial site (HCG Manavata Cancer Centre, HCG-MCC). The approval letter is available at: http://ctri.nic.in/Clinicaltrials//WriteReadData/ethic/7396280324DCGLEthicsCommitteeLetter.pdf http://ctri.nic.in/Clinicaltrials//WriteReadData/ethic/79159917MCRIEthicsCommitteeLetter.pdf

The trial was conducted in accordance with ethical guidelines and the Declaration of Helsinki. All study participants were previously counselled regarding study objectives as well as potential benefits and potential risks of the study. Written informed consent was obtained from all enrolled participants prior to enrollment. All study participants consented for publication of deidentified biological data. The present manuscript does not contain any personal or identifiable information or data of any participant. All authors have consented to publication of this manuscript and the data.

## Notes

**Research Support** This study did not receive any public or governmental funding or support. The study was wholly supported by the Sponsor, Datar Cancer Genetics (Pvt) Ltd.

### Competing Interest Statement

Rajnish Nagarkar has no competing interests; Darshana Patil, Dadasaheb Akolkar, Navin Srivastava, Vineet Datta, Sanket Patil, Sachin Apurwa, Ajay Srinivasan, are employees of the Study Sponsor; Rajan Datar is the founder of the Study Sponsor.

### Clinical Trial

CTRI/2019/02/017548

### Funding Statement

No external funding was obtained for this study. The entire study was funded by the Study Sponsor (DCG).

### Author Declarations

Ethical Considerations The Liquid Impact Prospective Study was approved by institutional review boards and ethics committees of the study sponsor (Datar Cancer Genetics, DCG) as well as clinical trial site (HCG Manavata Cancer Centre, HCG-MCC). The approval letter is available at: http://ctri.nic.in/Clinicaltrials//WriteReadData/ethic/7396280324DCGLEthicsCommitteeLetter.pdf http://ctri.nic.in/Clinicaltrials//WriteReadData/ethic/79159917MCRIEthicsCommitteeLetter.pdf The trial was conducted in accordance with ethical guidelines and the Declaration of Helsinki. All study participants were previously counselled regarding study objectives as well as potential benefits and potential risks of the study. Written informed consent was obtained from all enrolled participants prior to enrollment. All study participants consented for publication of deidentified biological data. The present manuscript does not contain any personal or identifiable information or data of any participant.

