## Supplementary methods for "Multi-Analyte Liquid Biopsies for Molecular Pathway Guided Personalized Treatment Selection in Advanced Refractory Cancers: A Clinical Utility Pilot Study"

**Blood Collection and Processing**

Approximately 10 mL peripheral blood was collected by venous puncture in each of the Cell-Free DNA BCT® and EDTA vacutainer tubes. Blood was stored and transported at 2-8°C. Plasma was separated by centrifugation at 3000× g for 20 min at 2-8°C, followed by 16000× g for 10 min at 20–25°C. Plasma from Cell-Free DNA BCT® tubes without hemolysis were processed for cell-free nucleic acid isolation.

**Cell-Free tumor DNA (ctDNA) Isolation**

Total ctDNA was purified from 2 mL plasma using a Circulating Nucleic Acid kit (QIAGEN, Germantown, USA) as per the manufacturer’s protocol. ctDNA was quantified using an HS DNA Qubit assay (Life Technologies, Carlsad, USA).

**Harvest of Viable Circulating Tumor Associated Cells (C-TACs)**

C-TACs were enriched and harvested from Peripheral Blood Mononuclear Cells (PBMCs) using a proprietary differentially cytotoxic medium as described previously ^1^. PBMCs were treated with a proprietary differentially cytotoxic medium for up to 100 hours at 37°C under 5% CO_2_, 4% O_2_. The medium induces cell death in normal (non-malignant) cells with functional apoptotic machinery while simultaneously conferring survival privilege on apoptosis-resistant cells of tumorigenic origin, i.e. Circulating Tumor Associated Cells (C-TACs; EpCAM+, PanCK+, CD45±) and their clusters. C-TACs include CTCs (EpCAM+, PanCK+, CD45-) as well as other cell types such as Tumor Associated Macrophages (TAMs) and Tumor Associated Fibroblasts (TAFs).

**DNA Mutation Profiling**

Tumor DNA (20 ng) was used for NGS library preparation via PCR-based Ampliseq target enrichment protocol. Libraries of 100 pmol were sequenced using Ion Proton (Thermo Fisher Scientific, Waltham, USA). Torrent Suite™ v5.2 (Thermo Fisher Scientific, Waltham, USA) software was used to perform primary analysis, including signal processing and base calling. Primary QC parameters were: minimum read length of 25 bases, read quality trimming of 17 QV, window size for quality trimming 30 bp. The processed sequenced data were aligned to the reference genome GRCh37/hg19 to generate Binary Alignment/Map (BAM) files. Sequencing data were considered for downstream analysis with coverage at ≥10,000× depth and >80% amplicons with at least 600 reads. The aligned data were analyzed using Torrent Variant Caller software with optimized parameters such as minimum allele frequency (0.003), minimum mapping quality (4), minimum coverage (600), down sample to coverage (10,000) and position bias (1). Reported somatic variants of >0.5% allele frequency (AF) were compared to the reference genome hg19. The Integrative Genomics Viewer (IGV) was used to visualize the read alignment and the presence of variants against the reference genome and to confirm the veracity of the variant calls by checking for possible strand biases and sequencing errors. All the germline variants found in the 1000 Genomes Project or The Exome Aggregation Consortium (ExAC) with a frequency of >0.1% were excluded. All somatic mutations were annotated, sorted and interpreted using COSMIC and/or TCGA data. Variants with <0.5% AF were confirmed orthogonally with digital droplet polymerase chain reaction (ddPCR, BioRad) using the rare mutation assay as per the manufacturer’s protocol.

A 50-gene NGS panel was used for ctDNA profiling and a 453-gene NGS panel was used for tumor DNA profiling to detect somatic hotspot mutations reported at high frequency in multiple cancer types as identified from TCGA, COSMIC, ICGC, MD Anderson Cancer Center and My Cancer Genome databases.

**Immunocytochemistry**

An aliquot of harvested C-TACs were fixed on slides with 4% paraformaldehyde (pH 6.9, 20 min). Cell permeabilization was achieved with 0.3% Triton-X 100 (15 min), followed by blocking with 3% BSA (30 min). Cells were treated with primary antibodies (60 min), washed with PBS (pH 7.4), incubated with secondary antibodies (60 min), washed with PBS and then incubated with 4’,6-Diamidino-2-phenylindole dihydrochloride (DAPI) in dark (15 min). All incubations were at ambient temperature (20°C – 25°C). Positive and negative cell line controls were also processed with each batch of samples. ICC slides were scanned by Cell Insight CX7 High-Content Screening (HCS) Platform (Thermo Fisher Scientific, USA) which enables nuclear size filters and calibration of intensity thresholds for individual fluorophore conjugated antibodies. The intensity of each antigen expression was compared to batch controls (reference cell lines). These precautions avoid or eliminate crosstalk in multiplexed analysis with different fluorophore conjugated antibodies. Aliquots of harvested cells were immunostained to determine status of EpCAM, Pan-CK and CD45 to confirm identity of C-TACs. Additional aliquots of harvested cells were then immunostained to determine status of theranostically relevant markers including VEGF, VEGFR, FGFR, PDGFR, EGFR, mTOR, AR, ER, HER2, TOPO and TUBB. Fluorescence imaging was performed on Cell Insight CX7 High-Content Screening Platform (ThermoFisher Scientific, USA).

**In vitro Chemo response Profiling**

The in vitro chemo response profiling (CRP) assay was designed ^2^ to evaluate the sensitivity of viable TDCs and CTACs to various anticancer agents (ACA). The test concentration for each ACA was based on reported peak plasma concentration at the recommended clinical dose and was preliminarily evaluated on SKBR3 (ATCC® HTB-30™), SW620 (ATCC® CCL-227™) and RCC 769-P (ATCC® CRL-1933™) cell lines and has been reported previously ^2^. Approximately 100 C-TACs/well were seeded into 96 well culture plates and incubated for 24 hours (37^°^C, 5% CO_2_, 4% O_2_). Viable cells were stained with Calcein AM and treated with ACAs. The plates were placed in the on-stage incubator of fluorescent microscope EVOS M7000 (Thermo Fisher Scientific) at 37^°^C with 5% CO_2_, 4% O_2_ and the wells imaged every 10 min for 12 h Extent of cell death was determined based on cell morphology changes and time required for fade out of live cell tracking dye.

All in vitro CRP assay plates included control wells (no drug) to determine baseline mortality as well as positive (known cytotoxicity) controls with SKBR3, SW620 or RCC 769-P cells. Cell death above 50% or >5KU after subtraction of baseline mortality in control wells were considered as an indication of drug efficacy. Drugs with higher activity were shortlisted for incorporation into patient regimens.

**Data Interpretation and Therapy Recommendation**

The process of data interpretation, integration to selection of final therapy was a semi-automated process which involved scoring followed by decision making by a multi-disciplinary tumor board. The process initially evaluated actionable gene variants, actionable gene expression (ICC) as well as in vitro chemosensitivity of viable C-TACs. The drug selection process prioritized selection of targeted and endocrine agents over cytotoxic anticancer agents for use in treatment regimens.

Targeted agents were initially shortlisted selected on the basis of gene variants (or features) that were known targets or the abundance of proteins detected by ICC. Gene variants with allele frequency ≥ 0.05% were deemed significant. All significant variants were queried against an in-house database to identify variations for which a targeted agent was indicated. ICC findings were deemed significant if the signal intensity of an antigen was ≥60% of the positive control. Patient-wise molecular indications were compiled along with the indications for targeted and endocrine agents.

A 50% threshold was established to discern drug resistance from responsiveness, i.e., drugs which yielded >50% mortality in C-TACs were considered for combination regimens. Drugs with >50% efficacy were evaluated using an in house database of combination regimens including those indicated in Standard of Care (SoC, e.g,, NCCN) as well as those combinations which have been previously evaluated in Phase I / II / III trials; the database prioritized safety over efficacy since the risk of unmanageable toxicity outweighed potential efficacy in a heavily pretreated population. Where multiple combinations with manageable toxicity profile could be generated from any of the selected drugs, these were further screened to prioritize the top 1 - 2 combinations where all or most drugs yielded >75% cell death by themselves.

In vitro chemosensitivity findings were integrated with the molecular findings and evaluated by a multi-disciplinary tumor board including the Principal Investigator who decided the final regimen based on patient fitness, prior treatments and anticipated toxicity profiles. Drug safety information was reviewed to generate a patient-wise list of expected AEs.

**SUPPLEMENTARY FIGURES**

**Supplementary Figure S1. Retrospective Study Cohort Dendrogram.** Graphical representation of sample source (commercial/research) and different components of the Multi-analyte Liquid Biopsy (MLB) analysis carried out in the retrospective study cohort.

**Supplementary Figure S2. Cancer-wise in vitro chemo response profile.** Graphical representation of percentage of samples in 33 cancer types demonstrating response to each of the 30 anticancer agents tested. Black bars represent SoC agents, while grey bars represent non-SoC agents in respective cancer types.

The 30 anticancer drugs tested include 5-Fluorouracil (5FU), Bleomycin (Ble), Cabazitaxel (Cab), Carboplatin (Car), Cisplatin (Cis), Cyclophosphamide (Cyc), Dacarbazine (Dac), Dactinomycin (Dct), Docetaxel (Doc), Doxorubicin (Dox), Epirubicin (Epi), Eribulin (Eri), Etoposide (Eto), Everolimus (Eve), Gemcitabine (Gem), Ifosfamide (Ifo), Irinotecan (Iri), Melphalan (Mel), Methotrexate (Met), Mitomycin (Mit), Mitoxantrone (Mix), Oxaliplatin (Oxa), Paclitaxel (Pac), Pemetrexed (Pem), Temozolomide (Tem), Topotecan (Top), Trabectedin (Tra), Vinblastine (Vib), Vincristine (Vic) and Vinorelbine (Vin).

**SUPPLEMENTARY TABLES**

**Supplementary Table S1. Demographics of Retrospective study cohort.** Details of gender stratification, median age and range, distribution of cancer types and organs and therapy status of the 1299 samples in the retrospective cohort.

**Supplementary Table S2. Cancer-wise in vitro chemo response profile.** Number and percentage of samples demonstrating sensitivities to the SoC and non-SoC anticancer agents tested in samples across different cancer types.

**Supplementary Table S3. Demographics of Prospective study cohort.** Details of gender stratification, median age and range, distribution of cancer types and organs and prior therapies of samples in the intent-to-treat (ITT) and evaluable subsets of the prospective cohort.

**Supplementary Table S4. Baseline Status.** FDG PET-CT scan based status of disease before initiation of treatment for patients in the prospective cohort.

**Supplementary Table S5. Prior Treatments.** Gender, age, cancer type and prior therapy details of patients in the prospective cohort.

**Supplementary Table S6. Treatments.** Distribution of cancer types across different treatment baskets such as mTOR, AGI, EGFR/ERBB2 and others.

**Supplementary Table S7. Outcomes.** Details of treatment outcomes, i.e., partial response, stable disease, median progression-free survival and median overall survival per treatment basket and for the overall study cohort.

**Supplementary Table S8. Patient wise AEs.** Basket-wise therapy-related adverse events in the ITT population with data across cancer type and grade.
