## Supplementary figures and images for "Multi-Analyte Liquid Biopsies for Molecular Pathway Guided Personalized Treatment Selection in Advanced Refractory Cancers: A Clinical Utility Pilot Study"

### Supplementary figure 1

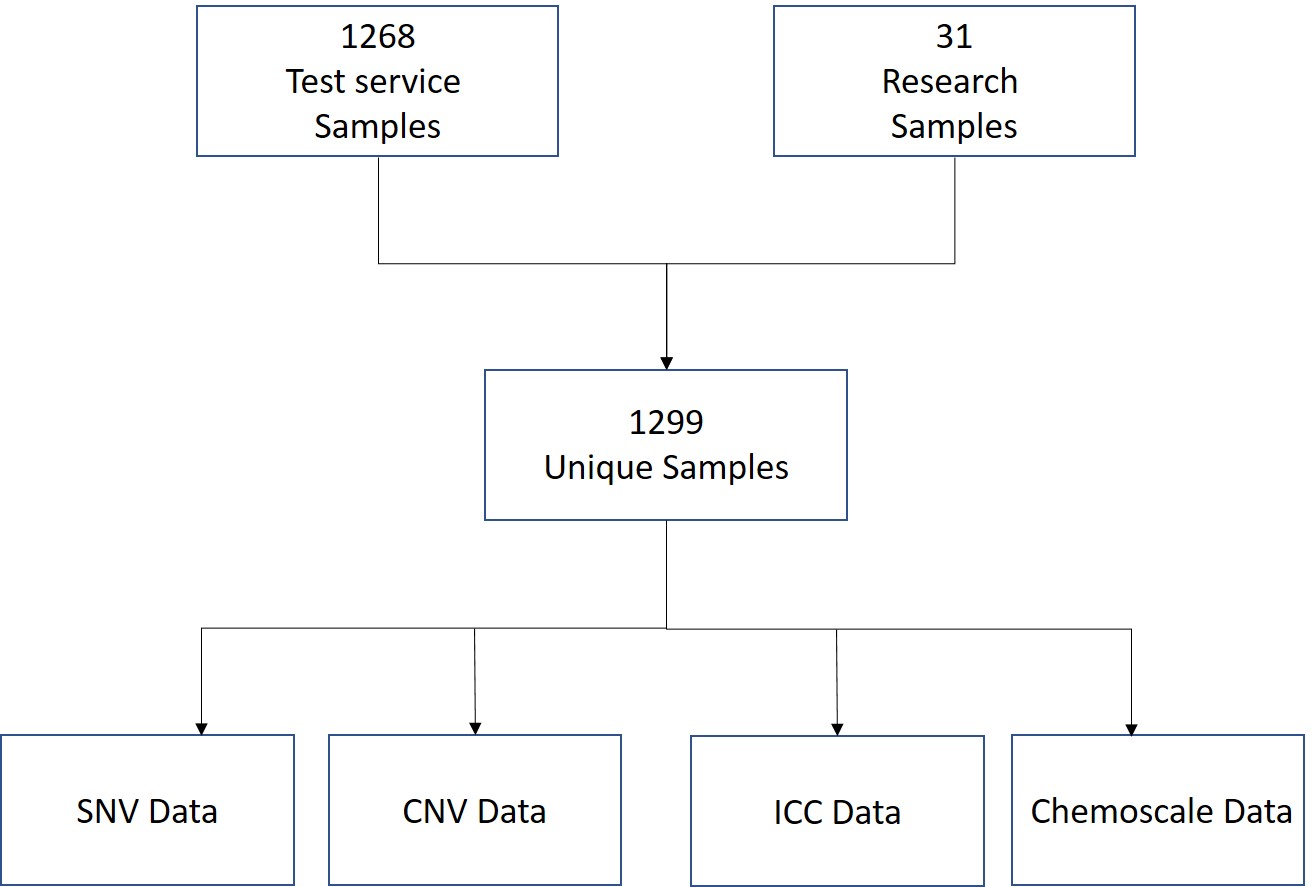

### Supplementary figure 2

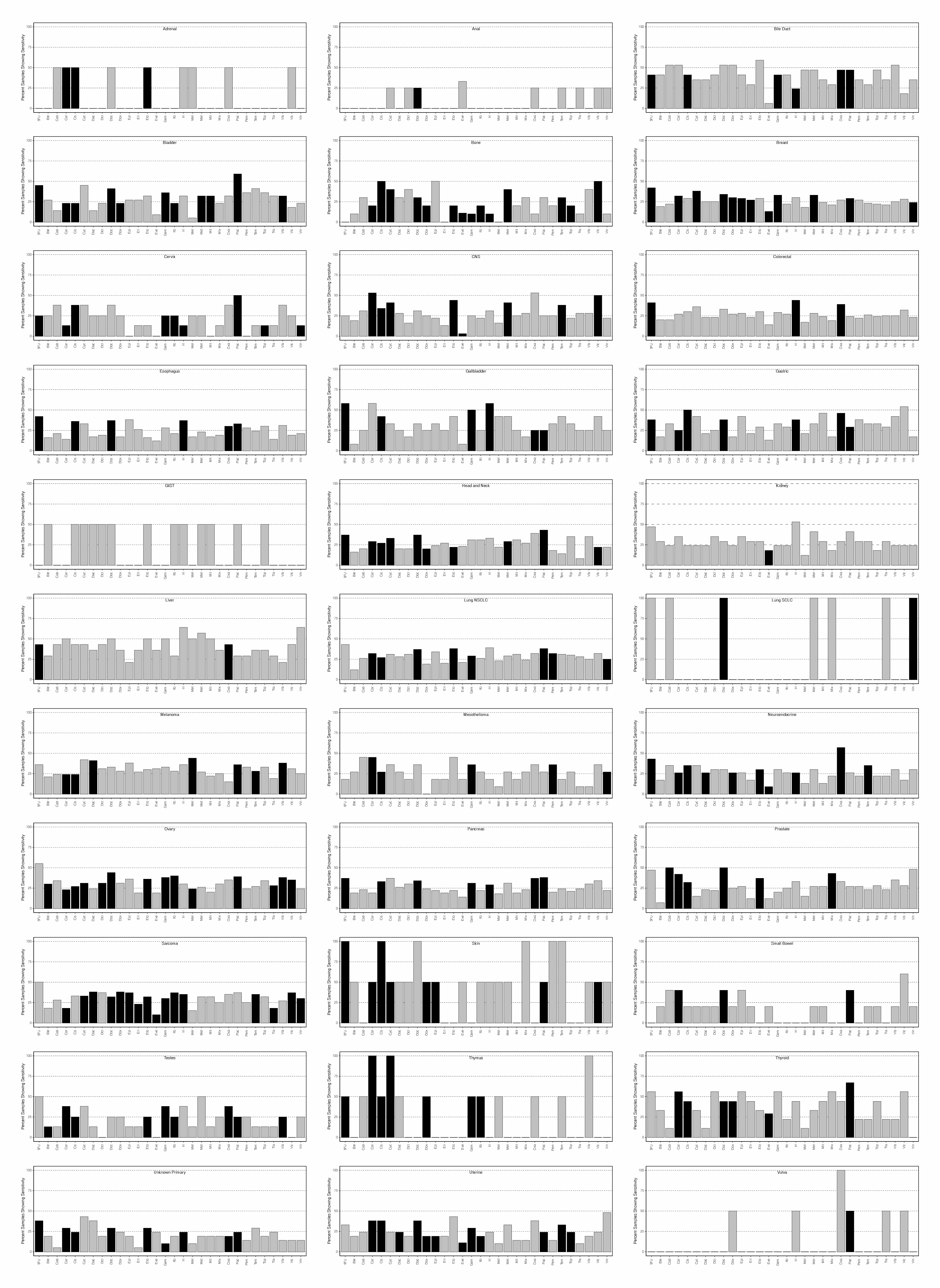
